# Characterizing the Progression of Pulmonary Edema Severity: Can Pairwise Comparisons in Radiology Reports Help?

**DOI:** 10.1101/2022.01.15.22269345

**Authors:** Stephanie M. Hu, Steven Horng, Seth J. Berkowitz, Ruizhi Liao, Rahul G. Krishnan, Li-wei H. Lehman, Roger G. Mark

**Affiliations:** Massachusetts Institute of Technology, Cambridge, MA, USA; Beth Israel Deaconess Medical Center, Harvard Medical School, Boston, MA, USA; Microsoft Research New England, Cambridge, MA, USA

## Abstract

Accurately assessing the severity of pulmonary edema is critical for making treatment decisions in congestive heart failure patients. However, the current scale for quantifying pulmonary edema based on chest radiographs does not have well-characterized severity levels, with substantial inter-radiologist disagreement. In this study, we investigate whether comparisons documented in radiology reports can provide accurate characterizations of pulmonary edema progression. We propose a rules-based natural language processing approach to assess the change in a patient’s pulmonary edema status (e.g. better, worse, no change) by performing pairwise comparisons of consecutive radiology reports, using regular expressions and heuristics derived from clinical knowledge. Evaluated against ground-truth labels from expert radiologists, our labeler extracts comparisons describing the progression of pulmonary edema with 0.875 precision and 0.891 recall. We also demonstrate the potential utility of comparison labels in providing additional fine-grained information over noisier labels produced by models that directly estimate severity level.

## Introduction

Pulmonary edema is a condition in which fluid accumulates in the lungs, making it difficult to breathe and ultimately leading to respiratory failure if treated improperly.^1^ It is often diagnosed using chest radiographs, which are interpreted by radiologists in a radiology report. Rather than the mere presence or absence of pulmonary edema, radiologists often assess the severity of the condition, which is critical in allowing clinicians to make better treatment decisions based on quantitative phenotyping of patient status.^2^ This is particularly important for congestive heart failure (CHF) patients, who demonstrate heterogeneous responses to treatment.^3^Unfortunately, accurate grading of pulmonary edema severity is a challenging task.^4^

The underlying physiology of pulmonary edema should be considered a continuous variable, but since humans are inaccurate at estimating continuous values, they instead use a discrete scale to rank the degree of the condition.^5,6^ While the current approach to quantifying pulmonary edema aligns with traditional practices in risk stratification, it also possesses shortcomings due to the difficulty in accurately assessing severity. To address this issue, we propose a rules-based natural language processing (NLP) algorithm to automatically extract comparison labels between consecutive intra-patient radiology reports in a series, as a way to provide more fine-grained and reliable information about pulmonary edema status.

The severity scale typically used by radiologists ranges from 0 to 3, with 0 indicating no pulmonary edema and 3 indicating the most severe level of pulmonary edema.^7,8^ In practice, however, the boundaries between these bins are not well defined and do not generalize effectively across patients. Furthermore, the bins often overlap between different radiologist interpretations, and there exists no consensus for objectively characterizing each bin (Figure 1). This makes it difficult to assign reliable severity labels to a patient’s pulmonary edema status and effectively inform their treatment plan. As a result, evaluation and treatment of pulmonary edema remains largely opinion-based.^9^

**Figure 1:**
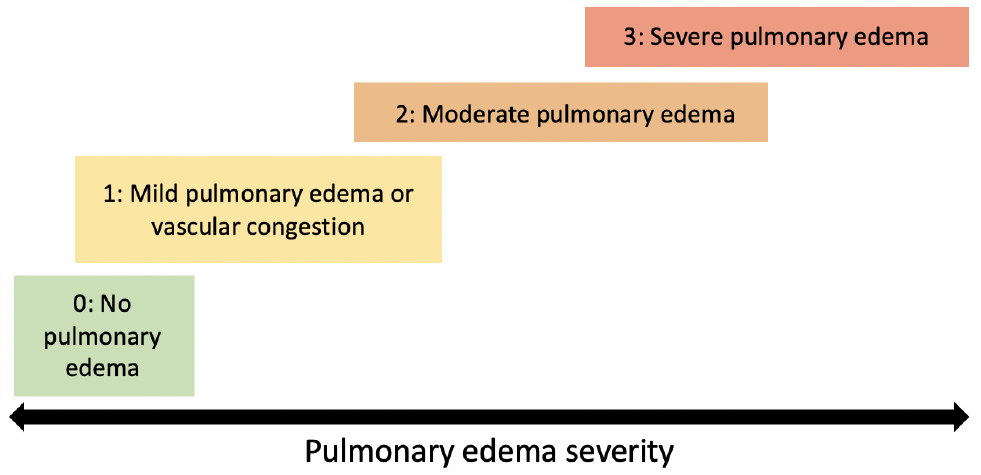
Schematic illustrating the overlap in pulmonary edema severity classifications.

To eliminate the subjectivity of human judgement, previous research developed a computer vision model for assigning severity scores to pulmonary edema patients based on chest radiographs.^10^ This effort follows a broader trend of using machine learning to improve the accuracy of radiograph interpretation and to assist with clinical decision making. The caveat is that most radiological data is unlabeled, which makes it difficult to train robust and reliable models. To address this challenge, researchers have tried using NLP techniques on radiology reports to extract labels for the associated radiographs.^10–12^ Irvin et al. released a large dataset of chest radiographs labeled by an automatic labeler that outperformed the current state-of-the-art NIH model.^11^ They developed a rules-based labeler to extract 14 different observations from radiology reports associated with chest X-rays and validated a subset of those labels using manual expert comparisons. Bustos et al. expanded upon this work by employing a recurrent neural network model with attention to label 174 different radiographic findings on their set of chest radiographs and associated reports.^12^

To specifically assess the severity of pulmonary edema, Liao et al. developed a program that employs keyword-matching to automatically extract pulmonary edema severity labels from radiology reports. These severity labels are subsequently used as “ground truth” for training a computer vision model to predict the severity of pulmonary edema from chest radiographs.^10^ However, this keyword-matching approach makes the assumption that the radiologist who interpreted the associated radiograph correctly quantified the status of pulmonary edema in the image, a task that has been proven difficult even for experts.^4^

While the severity score documented in the radiology report may not be accurate, there are other pieces of information that may be more reliable. In particular, when interpreting a radiograph, radiologists often make comparisons between the current radiograph and the previous radiograph in the series. These comparisons describe how radiologic features such as fluid status have changed, improved, or worsened over time. Since making comparisons is a task that is easier than estimating values on a scale, we propose using comparison labels to extract more granular information about pulmonary edema status. From these comparison labels, it may be possible to derive a continuous-valued scale for quantifying pulmonary edema that is more accurate and sensitive to the different levels of severity than the discrete system that is currently used. Indeed, there exist algorithms for constructing approximate rankings from pairwise comparisons that could be employed in future extensions of this work.^13,14^

Here we present a rules-based NLP approach for automatically assigning comparison labels to radiology reports that document changes in pulmonary edema status. The comparison labels are used to derive comparisons between pairs of consecutive reports, which can be used for a number of applications. The results presented here can assist researchers in developing more accurately labeled datasets for modeling, better inform radiologists trying to understand the characteristics of the different pulmonary edema severity levels, and help clinicians explore a continuous scale for severity grading. In turn, these outcomes have important implications for designing effective treatment plans for pulmonary edema and developing reliable tools for clinical decision-making.

### Dataset

In this study, we used radiology reports from the MIMIC-CXR database, which contains 369,071 chest radiographs and 222,856 associated radiology reports from 64,552 patients collected as part of routine clinical care at the Beth Israel Deaconess Medical Center.^15^ Since the same keywords can imply different clinical findings depending on disease context, we limited our cohort to CHF patients to reduce keyword confounding as in Horng et al.^16^ The average number of chest radiographs taken per CHF patient during a single hospital stay was 13.78 (compared to 5.43 for a non-CHF patient), making it possible to generate multiple pairwise comparisons for a given patient.

We further filtered our dataset to include only consecutive radiology reports. Two radiology reports were defined as *consecutive* if the associated radiographs were acquired within 48 hours of each other, and if no other radiographs were performed in between. In total, we used 7,141 radiology reports comprising 4,896 pairs across 1,114 patients in our study. Reports that were both preceded and followed by another report were included in two distinct pairs.

#### Comparison Labels

Given a pair of consecutive radiology reports *r* and *r*^*′*^ written at time *t* and *t*^*′*^, respectively, where *t*^*′*^ *> t*, we define a *comparison* to be any description of change on the patient’s pulmonary edema state between time *t* and *t*^*′*^. This change is captured in the text of *r*^*′*^, so the comparison label identified for the pair (*r, r*^*′*^) is simply the comparison label extracted from the document *r*^*′*^. Comparisons are either *worse, better*, or *no change*. We also used a *no comparison* label for documents that contained no explicit comparisons about pulmonary edema in the text. In our study, we assumed that any comparisons described in a given radiology report were made relative to the report dated immediately before it.

#### Training Set

Our labeler was developed using a training set of 257 radiology reports across 34 patients, where each radiology report was involved in one or two consecutive pairs. Because interpreting textual information is a straightforward task, we worked with a single radiologist to construct this “gold standard” dataset and received additional feedback from a second radiologist and a domain expert. We assigned comparison labels at the sentence level for sentences extracted from the “Findings” and “Impressions” sections of the radiology reports. These sentences were first identified as being relevant or not relevant to describing pulmonary edema status, and the relevant sentences were further assigned a comparison label capturing the change in severity of the condition. Characteristics of these sentences are detailed in Table 1. The 272 sentences that were considered to be relevant to pulmonary edema status yielded the following distribution of comparison classes: 36 worse, 40 better, 88 no change, and 108 no comparison.

**Table 1:** Characteristics of sentences in the training dataset. ^*\**^Observations were obtained by applying the CheXpert labeler^11^ to individual sentences.

|  |  |
| --- | --- |
| Total number of sentences extracted | 1,492 |
| Average sentence length (word count) | 10.14 |
| Median sentence length (word count) | 9 |
| Total number of sentences relevant to pulmonary edema | 272 |
| Average number of observations mentioned per sentence* | 1.34 |
| Average number of observations mentioned per relevant sentence* | 1.54 |

#### Testing Set

To evaluate the performance of our labeler in constructing pairwise comparisons at the document level, we randomly selected 101 pairs of consecutive radiology reports that had been labeled as one of *worse, better*, or *no change* by the labeler. None of the documents in this set overlapped with the training set. A board-certified radiologist, blinded from the results of the labeler, then provided manual comparison labels for these pairs at the document level only. Of the 101 pairs, the radiologist provided the following distribution of comparison labels: 24 worse, 26 better, 49 no change, and 2 no comparison. The 2 pairs of reports with the *no comparison* label were excluded from analysis, yielding a final testing set size of 99.

### Pairwise Comparison Labeler

An automatic rules-based labeler was developed for assigning pairwise comparison labels to consecutive radiology reports written in free text. The labeler consists of three main stages: 1) identifying sentences relevant to pulmonary edema, 2) identifying comparisons in individual sentences, and 3) constructing document-level pairwise comparison labels from sentence-level labels. The workflow is illustrated in Figure 2.

**Figure 2:**
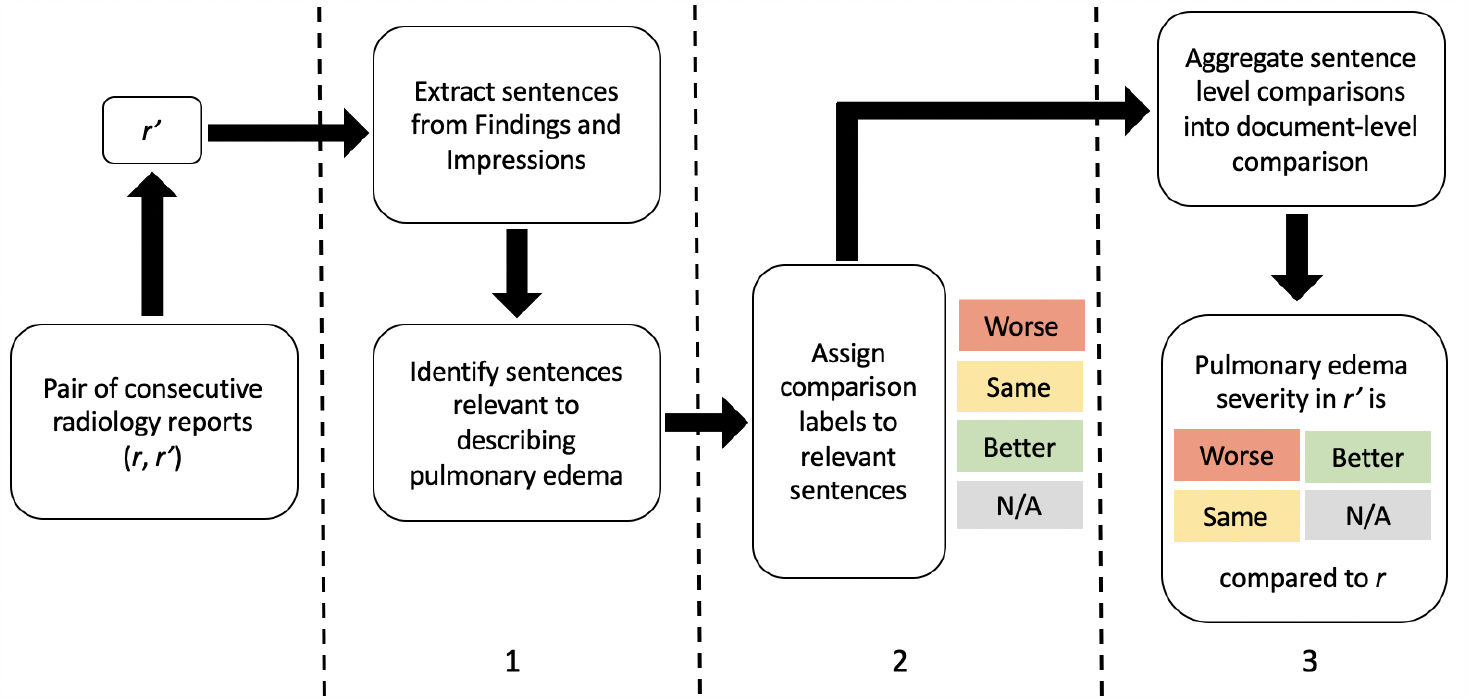
Labeler pipeline in three main stages.

### Identifying Relevance

The first step in our labeler is to extract individual sentences from the “Findings” and “Impressions” sections of the radiology reports, which generally capture the main observations, interpretations, and comparisons presented in the radiograph. A combination of rules-based methods is then used to identify the relevance of individual sentences in describing pulmonary edema status. For each of the methods discussed below, the output is a 1 if there is a positive mention of the observation, 0 if there is a negative mention, and None if there is no mention.

#### CheXpert Labeler

The CheXpert labeler is a rule-based labeler that extracts 14 different observations from the free text of radiology reports, including pulmonary edema.^11^ Our labeler applies CheXpert to individual sentences and uses the returned *Edema* observation to identify pulmonary edema relevance. Of the remaining 13 observations, *Lung Lesion, Consolidation, Pneumonia, Atelectasis, Pleural Effusion*, and *Pleural Other* are aggregated into a single *Other Finding Mention* label as these conditions may share differential diagnoses with pulmonary edema. Mentions of *Lung Opacity* without specific modifiers indicative of pulmonary edema (bilateral, parenchymal, alveolar, patchy, pulmonary, perihilar) are also included under the *Other Finding Mention* label.

#### Regex Labelers

Regular expressions, or *regexes* are character sequences that define a search pattern. In addition to letters and numbers, various other operators can be used that have special functions. For example, the regular expression *opaci(ties* |*fied*| *fication)* will match any of the following words: *opacities, opacified*, and *opacification*. Our labeler uses three sets of regular expressions developed in our study that describe different findings of interest: pulmonary edema mention, related radiologic observations, and a general “no change” condition. We expanded on the keywords curated by Liao et al.^10^ and Irvin et al.^11^ and compiled a comprehensive list of regular expressions capturing the various ways that pulmonary edema can be described in a radiology report. We also created a list of regular expressions to identify radiologic findings that are related to, but not definitive for, the presence of pulmonary edema. These lists were developed using findings reported in the literature and under the guidance of a radiologist.^7,8^

To identify sentences that describe a generally unchanged condition between consecutive radiographs, we created a third set of regular expressions that describe how a “no change” finding might be reported. It is important for our labeler to to consider these sentences because if a radiograph shows no change in patient status compared to the previous radiograph, the radiologist may not specifically document “no change in pulmonary edema” but rather more generally, “no change from prior.” Thus, even though pulmonary edema is not explicitly mentioned, the sentence is implicitly relevant and should be captured.

#### Aggregate Labeler

The output of the CheXpert and regex labelers described previously are combined using a set of rules devised with input from a radiologist. The aggregate labeler assigns a binary indicator to each sentence indicating if it is relevant to pulmonary edema. If either the CheXpert labeler or regex labelers identifies a mention of pulmonary edema (positive or negative) in a given sentence, then the sentence is considered relevant. If there is a general “no change” finding, then the sentence is also considered relevant. Sentences that mention related radiograph findings are only considered relevant if they do not contain mentions of other findings that could represent a differential diagnosis.

### Identifying Comparisons

In the second stage of our labeler, only sentences that are considered relevant to pulmonary edema are further assigned a comparison label. Our labeler uses a rules-based approach to assign one of four classes (*better, worse, no change*, or *no comparison*), using a set of regular expressions developed in our study to capture the directional change for each of the categories. The presence of a comparison phrase from one of the *better, worse* and *no change* categories was used to assign the appropriate comparison label. Absence of any comparisons results in a *no comparison* label.

#### Search Radius

While each sentence mentions only 1.54 observations on average (Table 1), some sentences contain descriptions for multiple observations. In order to increase the likelihood that a comparison phrase specifically describes pulmonary edema, we introduced a search radius parameter *p* in the labeler. Sentences are only assigned a comparison label if the comparison phrase is found within *p* words of an expression describing pulmonary edema, where *p* was determined experimentally.

#### Negation

Once comparison phrases are identified, the labeler uses the Negex library to determine whether the comparison is a negative or positive mention. If it is a negative mention (e.g. “*no improvement* in pulmonary edema”), then a *no change* comparison label is assigned to the sentence; otherwise, the original label is kept.

### Constructing Pairwise Comparisons

The third stage of our labeler aggregates sentence-level labels for a given radiology report into a single document-level comparison label. Any sentences without a comparison label are ignored when forming this final label. If all relevant comparison-containing sentences in the report are assigned the same class, then the radiology report is also assigned to that class. In the case of discrepancies between sentence-level labels, our labeler uses a set of rules devised based on domain knowledge. After consulting with a radiologist, we opted to employ a “last-first” approach, in which the last sentence describing a change in pulmonary edema status is given the highest precedent in determining the label of the entire radiology report. If there are no sentences explicitly mentioning pulmonary edema, the last sentence describing a change in a related radiographic feature is used as the document-level label. Reports that do not mention pulmonary edema but contain a general “no change” finding are assigned to the *no change* class. In a given pair of consecutive radiology reports (*r, r*), the pairwise comparison label is taken from *r*.

### Sample Case Study

Figure 3 walks through an example that illustrates how our labeler would classify a given radiology report into one of the four comparison classes. This comparison label describes the change observed in the associated radiograph relative to the radiograph taken before this one. Using the rules described in earlier sections, the final document-level label for this report is *better*.

**Figure 3:**
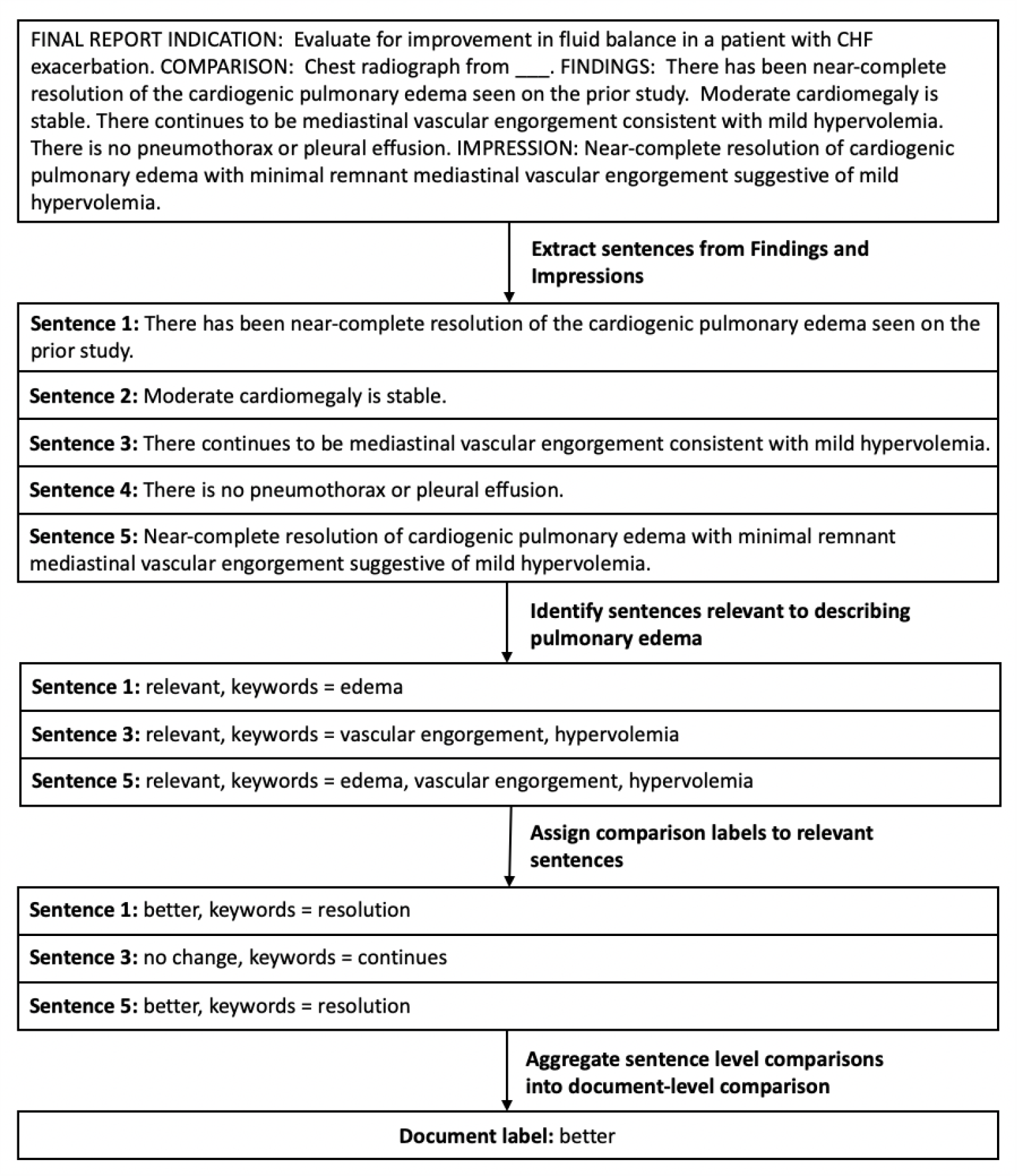
Simplified illustration of the labeling pipeline applied to a sample report.

### Performance on Training Set

We evaluated the performance of the first and second stages of our labeler against our training dataset, which contains radiology reports labeled at the sentence level. All 1,492 sentences in this set of radiologist reports were provided a relevance label by a radiologist, and the 272 sentences that were considered relevant to pulmonary edema were provided a comparison label.

#### Identifying Relevance

The goal of this first stage was to identify sentences relevant to characterizing pulmonary edema status in radiology reports. Because these sentences would later be aggregated to form document-level labels, we wanted to capture as many sentences as possible in this step. It was therefore more important to prioritize recall over precision. Our labeler achieved an accuracy of 98.8% on this task, with a precision of 0.96 and a recall of 0.98.

To demonstrate why targeting high recall makes sense in both this stage and the next stage of the labeler, consider the excerpts from sample radiology reports in Table 2. In the left report, sentences B and C are relevant to describing a change in pulmonary edema, while in the right report, sentence F is relevant. A classifier with perfect recall would label B, C, and F as relevant, but it may also label G as well. This is because B and G both describe a general change in patient status, and without context, it is difficult to ascertain whether this general change applies to pulmonary edema. However, the effect of (incorrectly) marking G as relevant is “undone” later in the pipeline when we aggregate all sentence-level labels into a single document-level label using the approach discussed previously. On the other hand, a classifier with perfect precision, at the expense of recall, would not label G as relevant to pulmonary edema, but it may also miss B as a result. This means that the only sentence labeled as relevant in the left report is C, which does not describe any comparison. Thus, at the end of the pipeline, the left report would ultimately be assigned a comparison label of *no comparison*, when it should actually be classified as *no change*.

**Table 2:** Examples of reports that are classified correctly by a labeler with high recall and low precision, but not by a labeler with high precision and low recall, since the left report would be incorrectly classified as *no comparison*.

|  |  |
| --- | --- |
| **A** The pleural effusions have slightly increased in extent. **B** Otherwise, no relevant changes seen. **C** Cardiomegaly, moderate-to-severe pulmonary edema present. **D** Rather extensive atelectasis at the lung bases. | **E** 1. Endotracheal tube in appropriate position, approximately 4 cm above the carina. **F** 2. Interval improvement in pulmonary vascular congestion. **G** 3. Otherwise, unchanged radiograph of the chest. |

#### Identifying Comparisons

We restricted our experiments and evaluation of this stage to the 272 sentences that were identified by a radiologist as relevant in determining pulmonary edema status. Metrics for individual classes were computed using a one-vs-all approach and are presented with the overall performance across classes in Table 3.

**Table 3:** Sentence-level results of comparison labeler on training set.

| Result | Accuracy | F1 | Precision | Recall |
| --- | --- | --- | --- | --- |
| Worse | 0.974 | 0.911 | 0.837 | 1.000 |
| No Change | 0.971 | 0.955 | 0.955 | 0.955 |
| Better | 0.992 | 0.974 | 1.000 | 0.950 |
| No Comparison | 0.952 | 0.939 | 0.962 | 0.917 |
| Overall | 0.967 | 0.946 | 0.949 | 0.945 |

We observe that the precision for the *worse* class is significantly lower than the precision for the *better* class, even though the approach that the labeler uses for assigning reports to either class is very similar, with the main difference being the expressions used for regular expression matching. This suggests that there may be a discrepancy in the way that results are reported for worsening versus improving pulmonary edema, and that additional analyses characterizing the structure of the radiology reports may be needed. As expected, the recall for the *no comparison* class is lower than the other classes, as we attempted to maximize, to a reasonable degree, the number of sentences assigned a comparison label in this stage of the labeler.

### Evaluation Against Radiologist Labels

The performance of our labeler in assigning pairwise comparisons to pairs of consecutive radiology reports was evaluated against a testing set of reports that were not used in the development of any stage of the labeling pipeline. The results are displayed in Table 4.

**Table 4:**
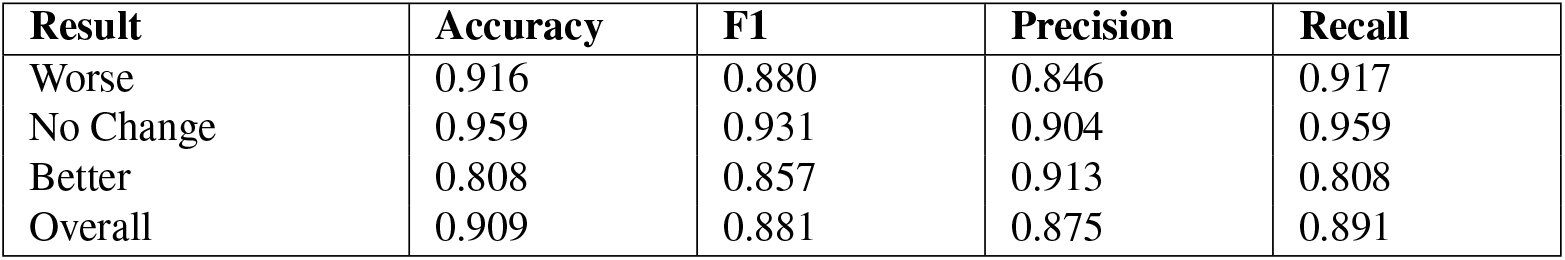
Document-level results of comparison labeler on testing set.

| Result | Accuracy | F1 | Precision | Recall |
| --- | --- | --- | --- | --- |
| Worse | 0.916 | 0.880 | 0.846 | 0.917 |
| No Change | 0.959 | 0.931 | 0.904 | 0.959 |
| Better | 0.808 | 0.857 | 0.913 | 0.808 |
| Overall | 0.909 | 0.881 | 0.875 | 0.891 |

Interestingly, even though the labeler performed best in terms of accuracy on the *better* class at the sentence level in the training set, it performs the worst on the testing set at the document level. The recall of the *better* class is also significantly lower. This suggests possible overfitting in the rules designed on the training set and/or an issue in the heuristics used to aggregate sentence-level comparisons into document-level labels.

Precision of the *worse* class is lowest at both the sentence and the document level, which again suggests that it should be handled differently than the *better* class by the labeler. One possible explanation for this low precision is that keywords such as *worse* and *greater* are commonly used in sentences describing differences between the left and right lungs (e.g. “again seen are bilateral lung opacities, asymmetric, right worse than left”). Using the rules we developed in our study, the labeler would classify such sentences as indicating worsening pulmonary edema. To better understand the types of misclassifications that our labeler is making, we present a confusion matrix on the testing set in Table 5.

**Table 5:** Distribution of class labels assigned by a radiologist versus the labeler developed in this study.

|  |  | Radiologist |  |  |
| --- | --- | --- | --- | --- |
|  |  | Worse | No change | Better |
| Labeler | Worse | 22 | 2 | 2 |
|  | No change | 0 | 47 | 3 |
|  | Better | 2 | 0 | 21 |

#### Existing Comparison Labelers

To the best of our knowledge, our work is the first attempt to automatically characterize pulmonary edema progression from radiology reports; however, there exist similar approaches for extracting change findings in other clinical conditions, such as brain tumors, using MRI reports.^17^ Even though Cheng et al. employed more complex machine learning techniques to discern tumor status, the performance of our labeler is similar to or improved over theirs for most comparison classes. This observation highlights a common trade-off in clinical informatics research between manually curated rules versus machine learning methods. The former requires domain expertise to help craft the rules for building the model while the latter requires domain expertise to label a large number of samples for training the model; ultimately, both approaches require a significant amount of time with domain experts. For some problems, such as the one addressed in our study, we argue that it is easier to manually create the rules, which have face validity to begin with, than to annotate a huge corpus of notes in hopes that a model will learn the rules. While more sophisticated computational methods may be necessary in some cases to discover patterns not obvious to human labelers, such as when describing tumor status in MRI reports, our results suggest that a simple rules-based model is largely sufficient to capture accurate comparison labels in other cases, such as for characterizing pulmonary edema progression in radiology reports.

#### Common Errors

While our labeler assigns correct labels to a majority of radiology reports, there are some cases in which it fails due to the presence of more complex sentences. In particular, when a sentence contains multiple observations close together, all with different modifiers, the labeler may select the wrong modifier to associate with the change in pulmonary edema status. For example, the sentence “Exam is otherwise remarkable for *improving* asymmetrical pulmonary edema and apparent *increase* in size of bilateral effusions” contains the comparison words “improving” and “increase” in close proximity to the pulmonary edema mention. In our algorithm, *worse* mentions take precedence over *better* and *no change*, so this sentence would be incorrectly classified as indicating worsening edema. On the other hand, there also exist sentences in which the same modifier applies to a number of observations. When that modifier lies too far away from the pulmonary edema mention, such as the keyword “unchanged” in the following example: “*Unchanged* evidence of moderate cardiomegaly, atelectasis, and overall mild-to-moderate *pulmonary edema*”, the labeler may not pick up on the presence of a comparison and incorrectly label the report as such.

### Comparison to Severity Labeler

To illustrate the advantage of extracting comparison labels from radiology reports over directly identifying the severity of pulmonary edema, we compared the output of our comparison labeler to the output of the keyword-based severity labeler developed by Liao et al.^10^ Because we wanted to see how pairwise severities compared to pairwise comparison in our evaluation, it was only meaningful to consider pairs of reports in which both reports had been assigned a severity label and in which the overall pair had been assigned a comparison label. In total, there were 243 such pairs. Table 6 summarizes the discrepancies in output labels between the two methods.

**Table 6:** Distribution of class labels assigned by comparison versus severity labelers. Note that the “comparison label” produced by the severity labeler is the directional change in severity scores between *r* and ^*′*^.

|  |  | Severity Labeler |  |  |
| --- | --- | --- | --- | --- |
|  |  | Worse | No change | Better |
| Comparison Labeler | Worse | 27 | 35 | 7 |
|  | No change | 15 | 92 | 24 |
|  | Better | 12 | 27 | 10 |

Nearly half of the observed discrepancies can be explained by the fact that the severity labeler only searches for the presence of keywords correlated to specific stages of pulmonary edema, without considering any modifiers. In other words, two reports that contain the same keyword will be labeled with the same severity score, even if that keyword was modified by comparison words such as *improving* or *worsening*. While this behavior is expected, it also shows that there are limitations to the severity labeler due to an inability to take into account the degree of change expressed. Table 7 highlights some examples.

**Table 7:** Excerpts from reports in which the severity labeler fails to take into account direction of change.

| Report $r$ | Report $r'$ | Predicted Severity (Both Reports) |
| --- | --- | --- |
| Since ... severe pulmonary edema is unchanged in severity | Over the last 12 hours, moderate-to-severe pulmonary edema has improved | 3 |
| As compared to prior radiograph several hrs earlier, pulmonary vascular congestion and mild interstitial edema are new | As compared to prior chest radiograph of ..., pulmonary vascular congestion and interstitial edema have nearly resolved | 2 |
| There is unchanged mild pulmonary vascular congestion | Previously seen mild vascular congestion and pulmonary edema has decreased | 1 |

Another possible source of discrepancy arises from the fact that humans are inherently better at making comparisons than assigning scores on a scale. As a result, if the radiologist who interpreted the radiograph reports the wrong score, then the severity labeler will pick up on the wrong score. Here is where the outputs of the comparison labeler may help refine the scores extracted by the severity labeler, as they are likely more reliable. Sample report pairs illustrating differences in inter-radiologist interpretations are provided in Table 8.

**Table 8:** Excerpts from reports in which the severity labeler suggests an incorrect change in pulmonary edema status between *r* and *r*^*′*^, likely due to differences in radiologist interpretation. The score assigned by the severity labeler is given in parentheses. The predicted comparison is based on the change in severity scores between *r* and *r*^*′*^ while the actual comparison is based on the comparison made in *r*^*′*^.

| Report $r$ | Report $r'$ | Predicted Comparison | Actual Comparison |
| --- | --- | --- | --- |
| Bilateral parenchymal opacities are present indicating moderate pulmonary edema. (3) | There is slight worsening of mild interstitial edema. (2) | Better | Worse |
| There is no confluent consolidation or frank pulmonary edema. (0) | Again seen is mild pulmonary vascular congestion, similar to the radiograph from ... (1) | Worse | No change |
| I do not believe there is pulmonary edema present. (0) | ...there is mild regression of pre-existing parenchymal opacities. (3) | Worse | Better |

### Comparison to Computer Vision Model

We also compared the results of our comparison labeler with the the computer vision model developed by Liao et al., which outputs severity labels for pulmonary edema based on radiograph images.^10^ Again, we only considered pairwise radiograph studies in which both images had a severity label and the pair of reports had a comparison label, and we analyzed the document-level comparison label from the comparison labeler and the signed difference between severities from the computer vision model. Out of the 2,404 pairs that were considered, 47% of them showed discrepancies between the two labelers (Table 9).

**Table 9:** Distribution of class labels assigned by comparison and severity labelers

|  |  | Computer Vision Model |  |  |
| --- | --- | --- | --- | --- |
|  |  | Worse | No change | Better |
| Comparison Labeler | Worse | 248 | 243 | 71 |
|  | No change | 249 | 729 | 275 |
|  | Better | 48 | 254 | 287 |

Although we did not perform a manual review to explain the discrepancies, we noticed that of all the pairs classified as *better* and *worse* by the comparison labeler, 43% of those pairs were indicated to have *no change* by the computer vision model. This supports the hypothesis that the computer vision model, which learns from the severity labels provided by the keyword labeler, doesn’t take into account more fine-grained differences between pulmonary edema status within the same severity class.

## Conclusion

In this study, we presented a rules-based approach for extracting comparisons on pulmonary edema status from pairwise radiology reports. Our labeler incorporates various NLP techniques with domain-specific clinical knowledge and achieves high performance on assessing directional change in pulmonary edema severity between consecutive radiology reports. These results may help clinicians gain more comprehensive insights into the pulmonary edema severity spectrum and better characterize the radiographic features that define each severity level. In the future, we would be interested in exploring more advanced algorithms to capture the complex cases that our current labeler misses, improving computer vision models by incorporating comparison labels during training, or constructing approximate rankings of pulmonary edema severity from pairwise comparisons.

## Data Availability

All data produced in the present study are available upon reasonable request to the authors.

## References

1. Mahdyoon H, Klein R, Eyler W, Lakier JB, Chakko S, Gheorghiade M. Radiographic pulmonary congestion in end-stage congestive heart failure. Am J Card. 1989 Mar 1;63(9):625–7.

2. Chakko S, Woska D, Martinez H, De Marchena E, Futterman L, Kessler KM, Myerburg RJ. Clinical, radiographic, and hemodynamic correlations in chronic congestive heart failure: conflicting results may lead to inappropriate care. Am J Med. 1991 Mar 1;90(1):353–9.

3. Francis GS, Cogswell R, Thenappan T. The heterogeneity of heart failure: will enhanced phenotyping be necessary for future clinical trial success? J Am Coll Cardiol. 2014 Oct 21;64(17):1775–6.

4. Hammon M, Danker P, Voit-Höhne HL, Sandmair M, Kammerer FJ, Uder M, Janka R. Improving diagnostic accuracy in assessing pulmonary edema on bedside chest radiographs using a standardized scoring approach. BMC Anesthesiol. 2014 Oct 18;14:94. PubMed PMID: 25364301.

5. Brady A, Laoide RÓ, McCarthy P, McDermott R. Discrepancy and Error in Radiology: Concepts, Causes and Consequences. Ulster Med J. 2012 Jan;81(1):3–9.

6. Warmbrod RJ. Reporting and Interpreting Scores Derived from Likert-Type Scales. J Agric Educ. 2014 Dec;55(5):30–47.

7. Cremers S, Bradshaw J, Herfkens F, editors. The Radiology Assistant: Heart Failure [Internet]. Radiological Society of the Netherlands; 2010 [cited 2020 Mar 22]. Available from: https://radiologyassistant.nl/chest/chest-x-ray/heart-failure.

8. Gluecker T, Capasso P, Schnyder P, Gudinchet F, Schaller MD, Revelly JP, Chiolero R, Vock P, Wicky S. Clinical and Radiologic Features of Pulmonary Edema. Radiographics. 1999 Nov 1;19(6):1507–31.

9. Chioncel O, Collins SP, Ambrosy AP, Gheorghiade M, Filippatos G. Pulmonary Oedema—Therapeutic Targets. Card Fail Rev. 2015 Apr 1;1(1):38–45.

10. Liao R, Rubin J, Lam G, Berkowitz S, Dalal S, Wells W, Horng S, Golland P. Semi-supervised learning for quantification of pulmonary edema in chest X-ray images. arixv;1902.10785 [Preprint]. 2019 [cited 2020 May 12]: [12 p.]. Available from: https://arxiv.org/abs/1902.10785.

11. Irvin J, Rajpurkar P, Ko M, Yu Y, Ciurea-Ilcus S, Chute C, et al. CheXpert: A large chest radiograph dataset with uncertainty labels and expert comparison. Proceedings of the AAAI Conference on Artificial Intelligence. 2019 Jul;33:590–7.

12. Bustos A, Pertusa A, Salinas JM, de la Iglesia-Vay’a M. Padchest: A large chest x-ray image dataset with multi-label annotated reports. 1901.07441 [Preprint]. 2019 [cited 2020 May 12]: [35 p.]. Available from: https://arxiv.org/abs/1901.07441.

13. Heckel R, Simchowitz M, Ramchandran K, Wainwright MJ, editors. Approximate ranking from pairwise comparisons. Proceedings of the Twenty-First International Conference on Artificial Intelligence and Statistics. PMLR: 2018 Apr;84:1057–66.

14. Shah NB, Wainwright MJ. Simple, Robust and Optimal Ranking from Pairwise Comparisons. J Mach Learn Res. 2018;18(199):1–38.

15. Johnson AEW, Pollard TJ, Berkowitz SJ, Greenbaum NR, Lungren MP, Deng CY, et al. MIMIC-CXR, a deidentified publicly available database of chest radiographs with free-text reports. Sci Data. 2019 Dec 12;6(317):1–8.

16. Horng S, Liao R, Wang X, Dalal S, Golland P, Berkowitz SJ. Deep learning to quantify pulmonary edema in chest radiographs. Radiology: Artificial Intelligence [Internet]. 2021 Jan 6 [cited 2021 Mar 5]: e190228. Available from: https://pubs.rsna.org/doi/abs/10.1148/ryai.2021190228.

17. Cheng LTE, Zheng J, Savova GK, Erickson BJ. Discerning Tumor Status from Unstructured MRI Reports—Completeness of Information in Existing Reports and Utility of Automated Natural Language Processing. J Digit Imaging. 2010 Apr;23(2):119–32.

